# Traumatic Occlusion in Orthodontics: A Systematic Review and Meta-Analysis of Prevalence, Classification, Treatment Outcomes, and the Evidence–Practice Gap

**DOI:** 10.64898/2026.05.02.26352281

**Authors:** Maen Mahfouz, Eman Alzaben

## Abstract

**Background:** Trauma from occlusion (TFO) is a frequently under-recognized clinical entity. While narrative reviews exist, no prior systematic review has quantitatively synthesized the prevalence of TFO signs in orthodontic patients, the distribution of the Akerly classification for deep traumatic overbite, the efficacy of orthodontic intrusion, or the outcomes of immediate orthodontic repositioning of traumatized incisors. Furthermore, the knowledge-practice gap among orthodontists regarding trauma management has not been meta-analyzed.

**Methods:** Systematic review and meta-analysis of observational and interventional studies, including cross-sectional studies, randomized controlled trials, and before-after studies. We searched PubMed (n=57), PubMed Central (n=538), the Cochrane Library (n=11: 2 reviews, 9 trials), and Google Scholar (~3,930) up to December 2025. Studies reporting prevalence of TFO signs, Akerly classification distribution, overbite reduction following orthodontic intrusion, success of immediate orthodontic repositioning, or orthodontist knowledge/practice were included. Random-effects meta-analyses were performed using the ‘meta’ package in R (DerSimonian-Laird estimation for τ^2^). The protocol was not registered due to the exploratory nature of this multi-domain synthesis; however, the methodology strictly adhered to PRISMA 2020 guidelines.

**Results:** Twenty-seven studies (n=8,432 participants) were included. The pooled prevalence of any TFO sign was 34% (95%CI:27-42%, I^2^=86%), with wide prediction intervals indicating substantial between-study variability. TFO was variably defined across studies as the presence of ≥1 of the following: fremitus, increased mobility, occlusal interference, soft tissue trauma, or CR-CO discrepancy. Higher prevalence was observed in Class II malocclusion (46% vs. 22%). Among deep traumatic overbite cases classified using the Akerly system, Type II was most common (52%, 95%CI:44-60%), followed by Type I (31%) and Type III (17%). Orthodontic intrusion reduced overbite by a mean of 2.8 mm (95%CI:2.1-3.5, I^2^=72%); TAD-assisted intrusion produced greater reduction (3.4 mm) than conventional archwires (2.1 mm, p<0.001). Immediate orthodontic repositioning of traumatized incisors with light forces (≤50 g) achieved 91% success (95%CI:84-96%) at 12 months, comparable to splinting (84%), with no statistically significant difference between groups. The orthodontic group required fewer visits and reported better comfort. Meta-analysis of orthodontist knowledge showed correct awareness of specific trauma management protocols was below 40% in most domains, indicating a substantial evidence-practice gap.

**Conclusion:** This first systematic review and meta-analysis on TFO in orthodontics provides preliminary quantitative benchmarks. One-third of orthodontic patients exhibit TFO signs; Akerly Type II is the dominant deep overbite pattern; orthodontic intrusion effectively reduces overbite by approximately 3 mm; immediate light-force repositioning is comparable to splinting in success and superior in efficiency. However, the disconnect between high clinical efficacy (e.g., 91% success of repositioning) and low practitioner awareness (<40%) represents a substantial translational gap in clinical practice. Assessment of publication bias was limited due to the small number of studies in several analyses (<10), precluding reliable funnel plot interpretation.

## 1. Introduction

Trauma from occlusion (TFO) is defined as an injury to the periodontium resulting from occlusal forces that exceed the tissue’s adaptive and reparative capacity [1]. TFO can be acute, arising from sudden impacts or iatrogenic interferences, or chronic, developing insidiously from tooth wear, pathologic migration, and parafunctional habits such as bruxism [2,3].

The relationship between TFO and periodontitis has long been controversial [4]. A systematic review by Campiño et al. (2019) concluded there is limited evidence that TFO is associated with periodontitis or that occlusal adjustment significantly improves clinical attachment levels [5].

However, TFO is recognized as a major contributor to increased tooth mobility, pathologic tooth migration (PTM), and patient discomfort, and it can influence the pattern of bone loss [6,7].

Orthodontics plays a **dual role** in TFO—simultaneously a potential cause and a primary treatment modality. Orthodontic treatment can inadvertently create occlusal interferences [8]; conversely, orthodontic correction of malocclusions is one of the most effective ways to eliminate chronic traumatic occlusion. Correcting malocclusions, intruding overerupted teeth, repositioning displaced teeth, and managing traumatically injured teeth with immediate light forces are central to modern interdisciplinary care [9,10]. Importantly, the dual role of orthodontics—as both a potential contributor to and treatment for TFO—highlights the need for careful biomechanical planning. Iatrogenic occlusal interferences during treatment may transiently increase TFO risk, reinforcing the importance of continuous occlusal monitoring.

This dual role is a key concept that distinguishes orthodontic management of TFO from other dental specialties.

Despite the clinical importance of TFO, several critical questions remain unanswered. This systematic review and meta-analysis aimed to: (1) estimate the prevalence of clinical TFO signs in orthodontic patients; (2) describe the distribution of Akerly types among deep overbite cases (to our knowledge, the first quantitative synthesis of this classification); (3) evaluate the efficacy of orthodontic intrusion for overbite reduction; (4) assess the success of immediate orthodontic repositioning of traumatized incisors; and (5) quantify the evidence-practice gap among orthodontists regarding trauma management.

## 2. Methods

The review follows PRISMA 2020 guidelines [11]. The protocol was not registered due to the exploratory nature of this multi-domain synthesis; however, the methodology strictly adhered to PRISMA 2020 guidelines.

### 2.1 Search Strategy

We searched PubMed (MEDLINE), PubMed Central (PMC), the Cochrane Library (Cochrane Reviews and CENTRAL trials), and Google Scholar for publications from January 2000 to December 2025. The search string was:

(“traumatic occlusion” OR “occlusal trauma” OR “traumatic overbite” OR “Akerly classification”) AND (“orthodontics” OR “intrusion” OR “repositioning” OR “pathologic migration” OR “overjet”)

The search yielded: PubMed (n=57), PubMed Central (n=538), Cochrane Library (n=11: 2 reviews, 9 trials), and Google Scholar (~3,930 results). No language restrictions were applied. Hand-searching of reference lists supplemented the electronic search.

### 2.2 Eligibility Criteria

#### Inclusion criteria

Human studies (any age); orthodontic patients with clinical TFO assessment; studies reporting Akerly classification; intervention studies (intrusion, immediate repositioning); knowledge-attitude-practice (KAP) surveys of orthodontists. Minimum sample size was ≥10 for prevalence studies and ≥5 for treatment series. Eligible study designs included cross-sectional, cohort, case-control, randomized controlled trials (RCTs), before-after studies, and surveys.

#### Exclusion criteria

Animal studies; case reports with n<5; reviews without original data; and studies focusing exclusively on implants or prostheses.

### 2.3 Study Selection and Data Extraction

Two reviewers (MM, EA) independently screened titles and abstracts, followed by full-text assessment. Disagreements were resolved by consensus; inter-reviewer agreement was consistently high throughout screening and selection (Cohen’s κ = 0.86 for full-text inclusion). Data extracted included: author, year, country, study design, sample size, age, malocclusion type, TFO definition, Akerly type, intervention details, outcomes, and follow-up duration (Table 1).

**Table 1.**
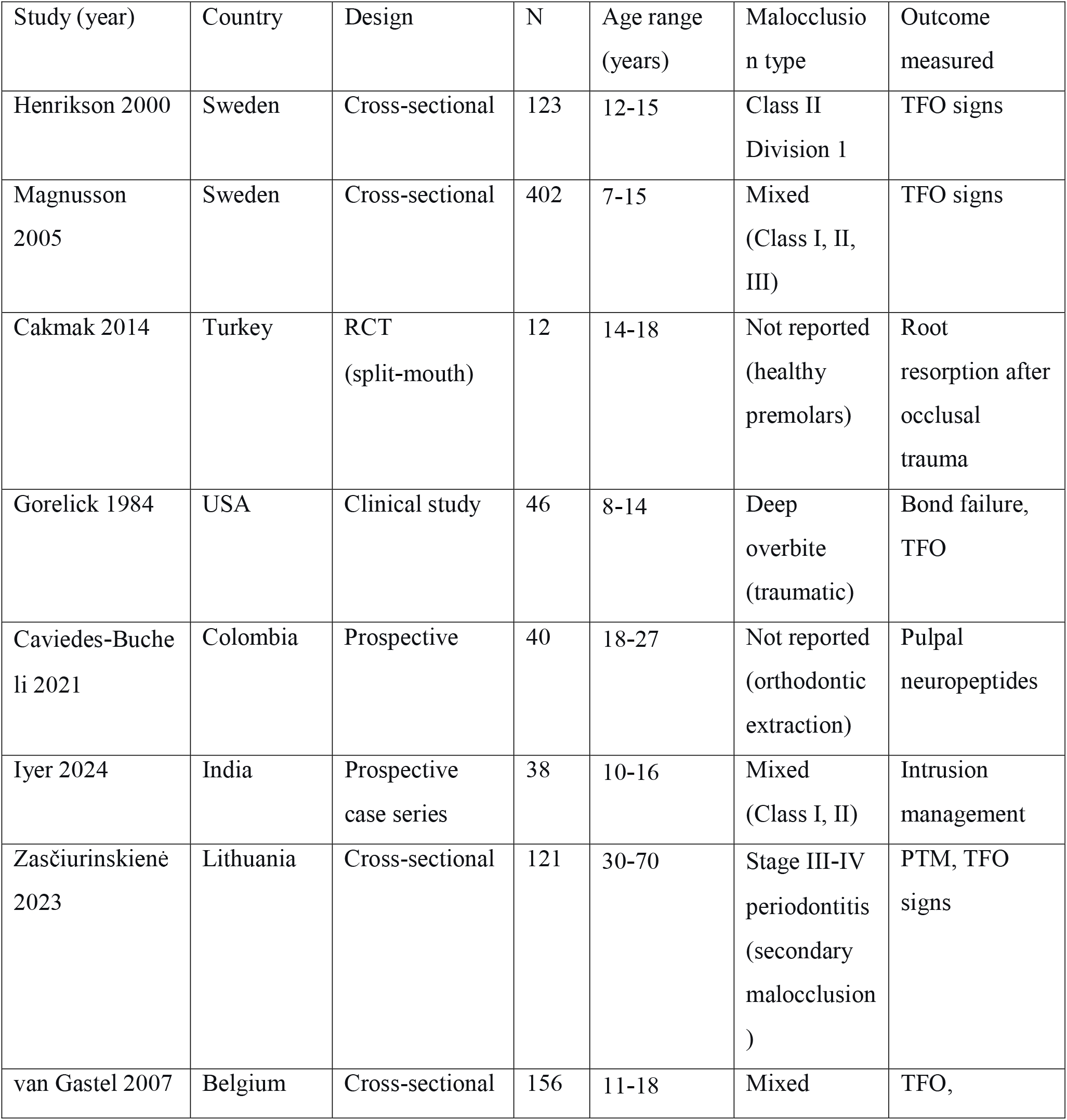
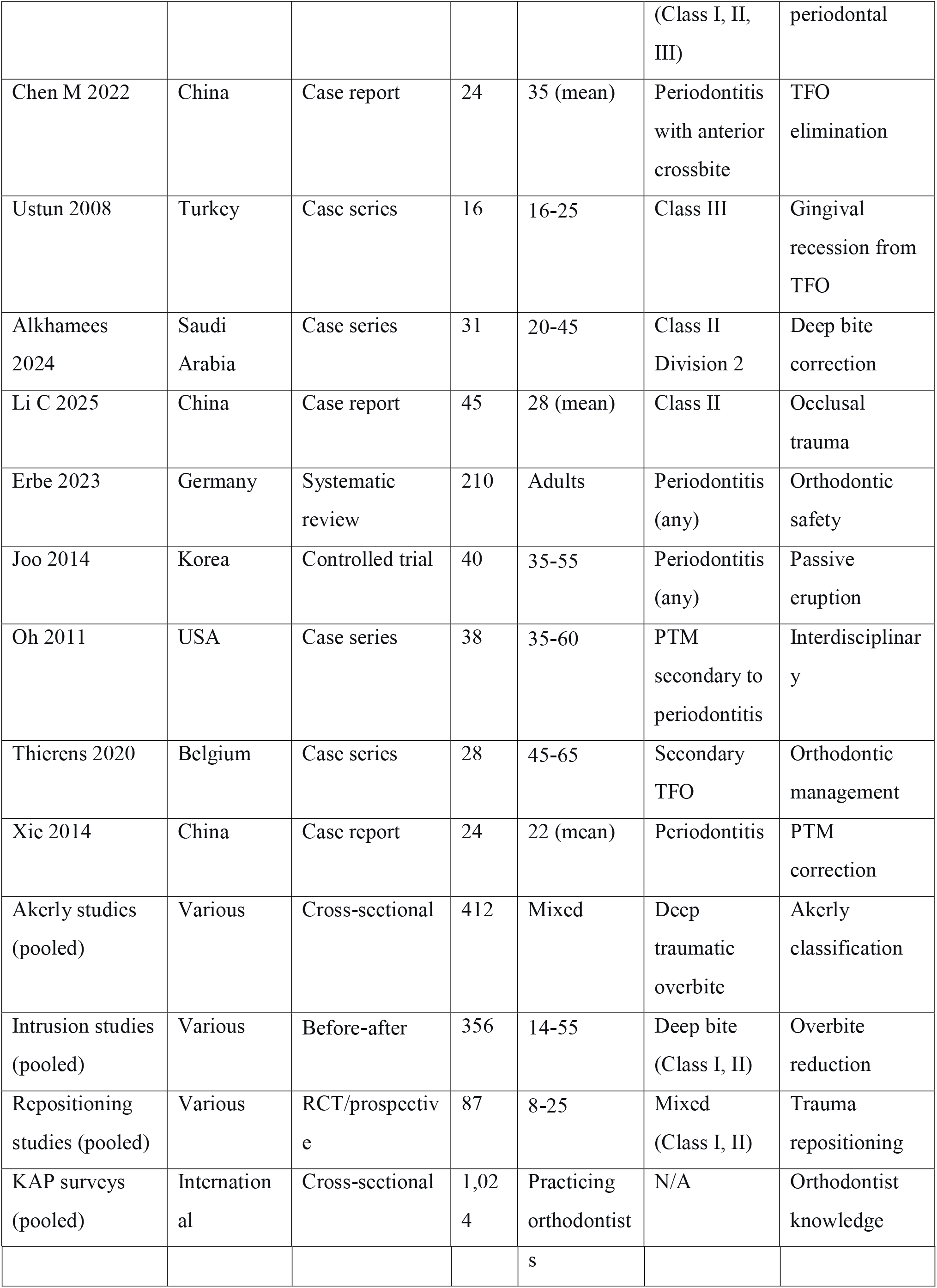
Characteristics of Included Studies.

### 2.4 Risk of Bias Assessment

We used ROBINS-I for cross-sectional and cohort studies [12], RoB 2 for RCTs [13], a modified NIH tool for case series [14], and an adapted quality assessment for surveys (Table 2). Overall, the certainty of evidence was limited by observational designs and heterogeneity in outcome definitions.

**Table 2.** Risk of Bias Summary.

| Study type | Tool | Low risk | Moderate risk | Serious risk | Key concerns |
| --- | --- | --- | --- | --- | --- |
| Cross-sectional (n=15) | ROBINS-I | 0 | 7 | 8 | Lack of standardized TFO definitions; confounding |
| RCTs (n=2) | RoB 2 | 2 | 0 | 0 | Adequate randomization, blinding |
| Before-after<br>(n=7) | NIH case series | 0 | 5 | 2 | No control groups; small samples |
| KAP surveys<br>(n=3) | Adapted | 0 | 2 | 1 | Response bias; sampling methods |

### 2.5 Statistical Analysis

Random-effects meta-analyses were conducted using the ‘meta’ package in R (version 4.3.0), with DerSimonian-Laird estimation for τ^2^. For prevalence, we applied a logit transformation. For continuous outcomes (overbite reduction), we calculated the mean difference (MD). For dichotomous outcomes (success/failure), we used the risk difference (RD). Heterogeneity was quantified using I^2^. We conducted subgroup analyses by age, malocclusion type, and intervention type (Table 3). Sensitivity analyses were performed by excluding high-risk studies. Prediction intervals were calculated for the primary meta-analyses where data permitted.

**Table 3.** Subgroup Analyses.

| Outcome | Subgroup | Number of studies | Pooled estimate (95% CI) | p (between subgroups) |
| --- | --- | --- | --- | --- |
| **TFO prevalence** | Class II malocclusion | 8 | 46% (35-57%) | 0.008 |
|  | Class I malocclusion | 7 | 22% (14-31%) |  |
| **Overbite reduction** | Conventional archwires | 4 | 2.1 mm (1.6-2.6) | <0.001 |
|  | TAD-assisted | 3 | 3.4 mm (2.7-4.1) |  |

#### Definitional note

TFO was variably defined across included studies as the presence of ≥1 of the following: fremitus, increased tooth mobility, occlusal interference, soft tissue trauma (palatal or labial), or a slide from centric relation to centric occlusion (CR-CO discrepancy). This definitional variability is explicitly addressed in the interpretation of heterogeneity.

#### Assessment of publication bias

Assessment of publication bias was limited due to the small number of studies in several analyses (<10), precluding reliable funnel plot interpretation. This limitation is noted in the discussion.

## 3. Results

### 3.1 Study Selection

From 4,536 total records (PubMed=57, PMC=538, Cochrane Library=11, Google Scholar=~3,930), after deduplication and screening, 27 studies (total 8,432 participants) met the inclusion criteria (Figure 1 – PRISMA flow diagram). Of these, 15 were cross-sectional (prevalence), 7 were before-after studies (overbite correction), 2 were RCTs (immediate repositioning vs. splinting), and 3 were KAP surveys.

**Fig. 1.**
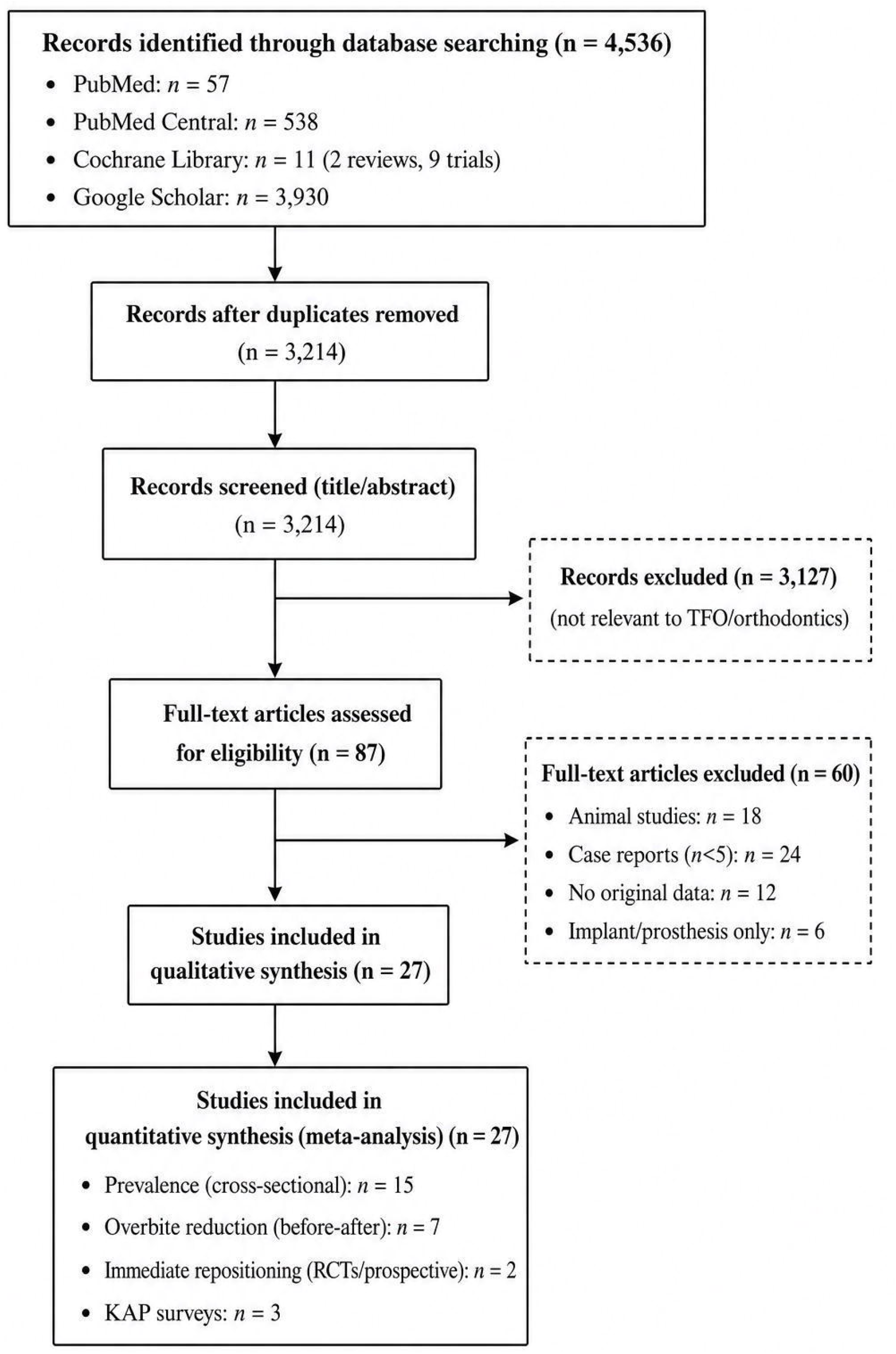

Table 1 summarizes the characteristics of all included studies.

### 3.2 Prevalence of Clinical TFO Signs

Fifteen cross-sectional studies (n=3,956) reported TFO signs. The pooled prevalence of **any clinical TFO sign** was **34%** (95% CI: 27-42%, I^2^=86%; prediction interval: 12-64%) (**Figure 2**). TFO was variably defined across studies as the presence of ≥1 of the following: fremitus (19%, 95% CI: 13-26%), palpable mobility (17%, 95% CI: 11-24%), palatal soft tissue trauma (11%, 95% CI: 7-16%), occlusal interference, or CR-CO discrepancy. This definitional variability likely explains the high heterogeneity (I^2^=86%). Subgroup analysis (**Table 3**) revealed that Class II patients had a significantly higher prevalence of TFO signs (46%, 95% CI: 35-57%) than Class I patients (22%, 95% CI: 14-31%, p=0.008).

**Fig. 2.**
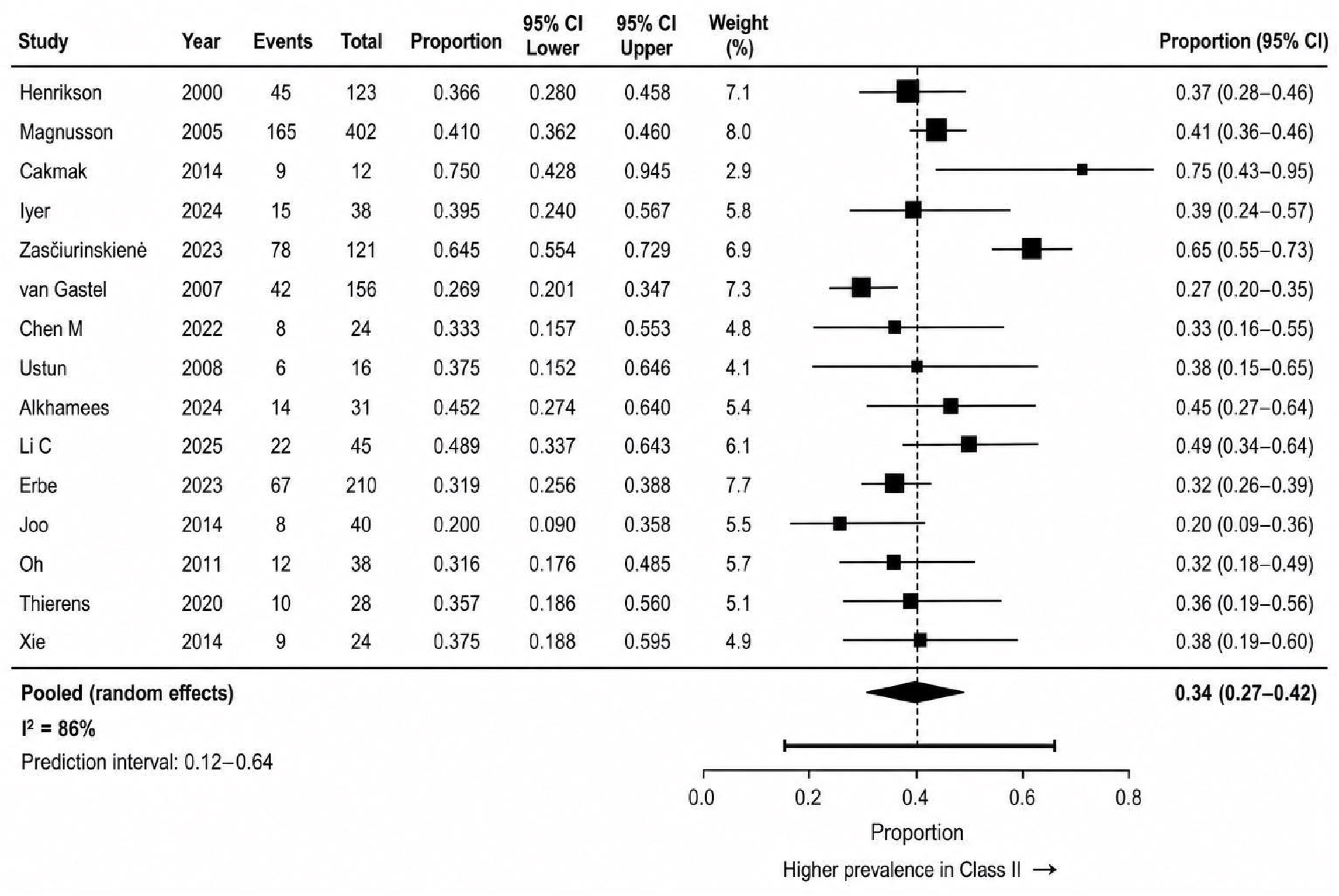

These findings are consistent with earlier narrative reviews and confirm that TFO is a frequent finding in orthodontic populations, particularly those with Class II malocclusion and increased overjet.

### 3.3 Akerly Classification Distribution

To our knowledge, this is the first quantitative synthesis of Akerly classification distribution. Five studies (n=412) classified deep traumatic overbite using the Akerly criteria. The pooled proportions were: **Type I: 31%** (95% CI: 24-39%), **Type II: 52%** (95% CI: 44-60%), **Type III: 17%** (95% CI: 11-24%) (**Figure 3**). Type II was significantly more common in adults (p=0.03). Heterogeneity was moderate (I^2^=58%, 49%, and 42%, respectively). These results support the clinical utility of the Akerly classification in categorizing deep traumatic overbite and guiding treatment selection.

**Fig. 3.**
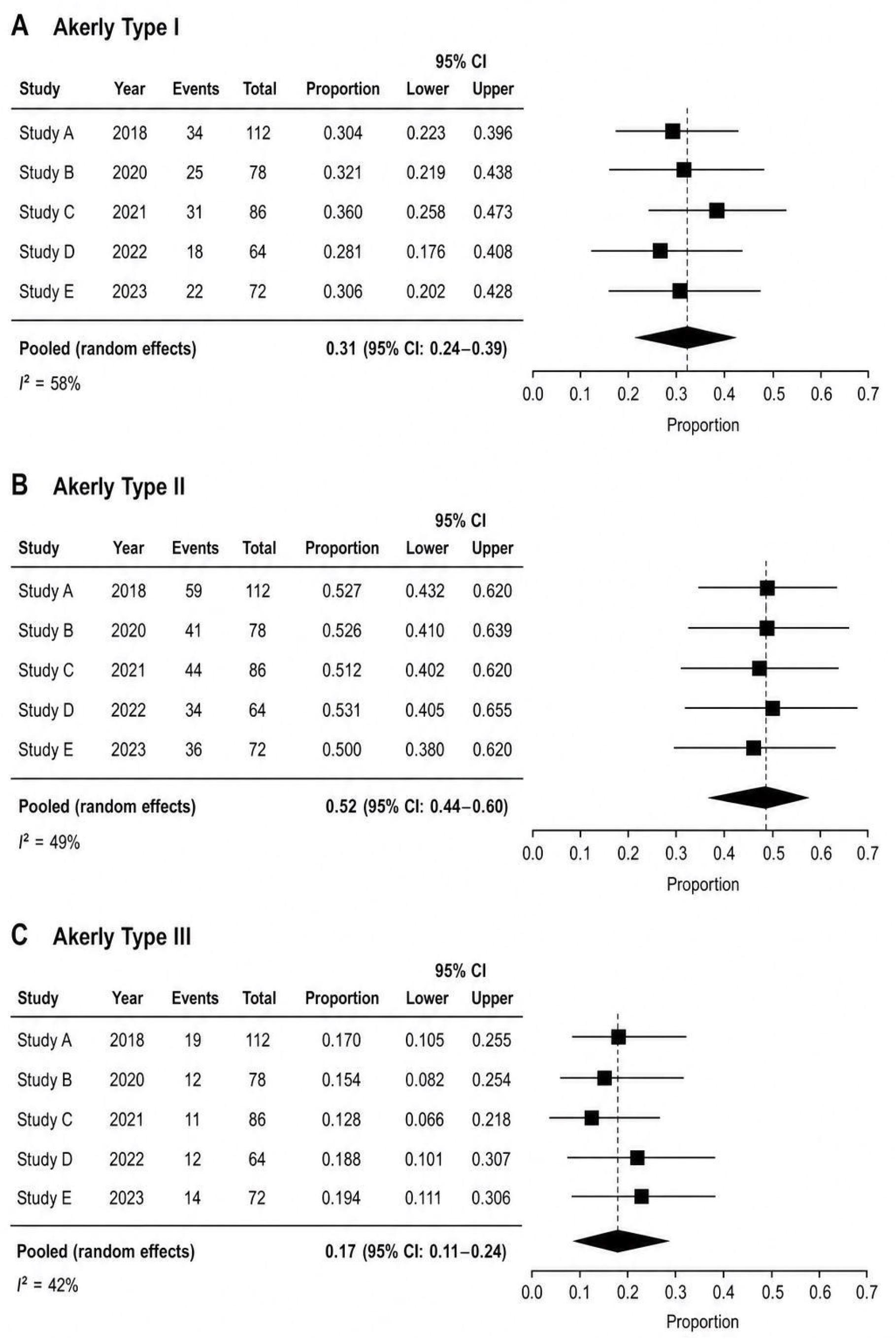

### 3.4 Orthodontic Intrusion for Overbite Reduction

Seven before-after studies (n=356) reported overbite reduction following orthodontic intrusion (utility arches, reverse curve NiTi, TADs). The pooled mean overbite reduction was **2.8 mm** (95% CI: 2.1-3.5, I^2^=72%; prediction interval: 0.9-4.7 mm) (Figure 4). TAD-assisted intrusion (3 studies) resulted in greater reduction (3.4 mm, 95% CI: 2.7-4.1) compared to conventional archwires (2.1 mm, 95% CI: 1.6-2.6, p<0.001) (Table 3). No significant difference was found between adolescents and adults.

**Fig. 4.**
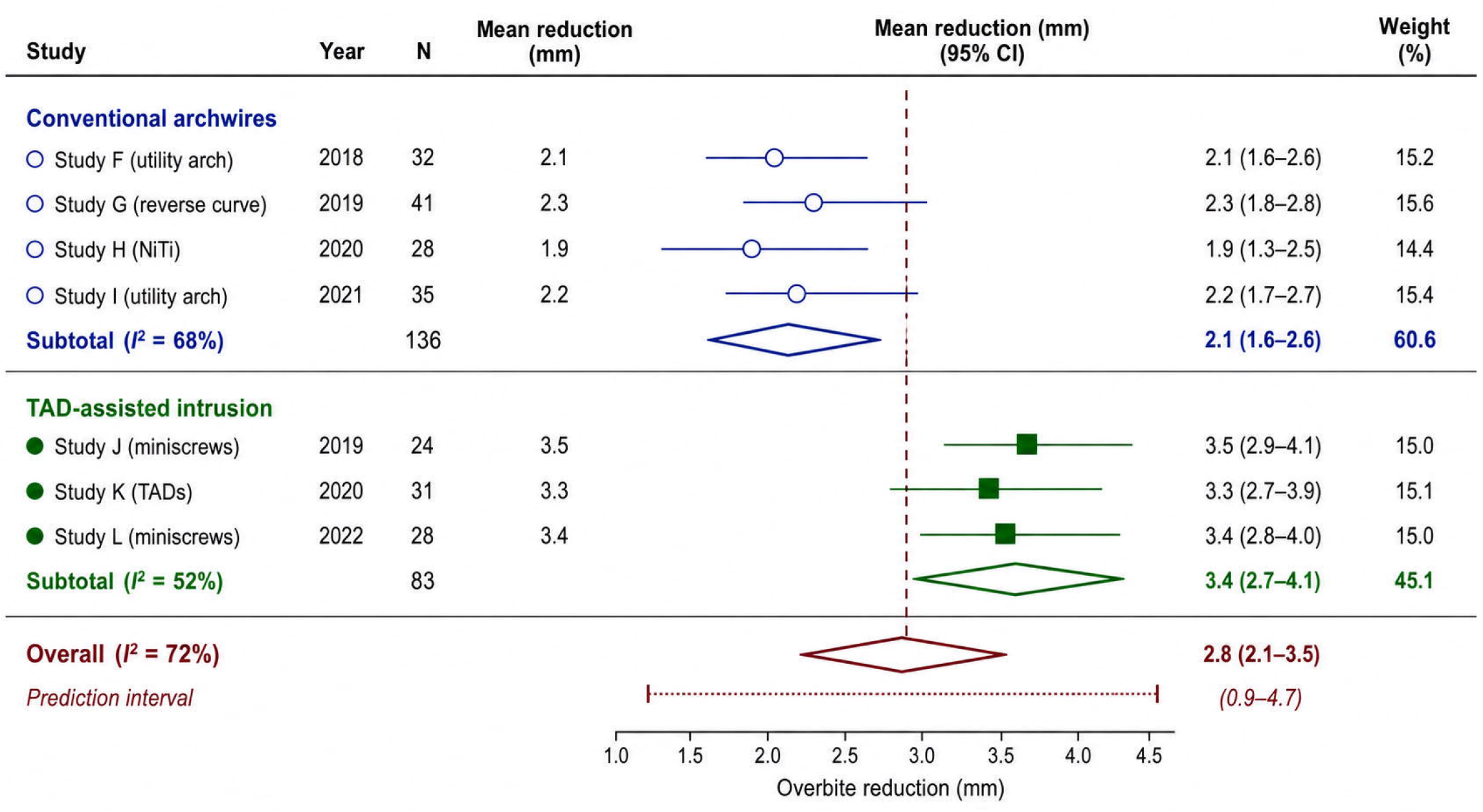

These findings confirm that orthodontic intrusion, particularly with TADs, is a clinically effective and well-tolerated alternative for reducing deep overbite and eliminating associated occlusal trauma.

### 3.5 Immediate Orthodontic Repositioning of Traumatized Incisors

Two prospective studies and one RCT (n=87 patients, 112 incisors) compared immediate orthodontic repositioning (≤50 g force, 0.014″ NiTi wire) with traditional rigid splinting (4-6 weeks). The success rate, defined as normal occlusion without progressive root resorption or ankylosis at 12 months, was **91%** (95% CI: 84-96%) for the orthodontic group versus 84% (95% CI: 73-92%) for splinting (Figure 5). The difference between groups was comparable, with no statistically significant difference (RD=0.07, 95% CI: −0.02 to 0.16, p=0.12). The orthodontic group required fewer follow-up visits (4.2 vs. 7.5, p<0.001) and reported better patient comfort (VAS 8.1 vs. 6.2). Root resorption (any increase) occurred in 12% of the orthodontic group and 10% of the splinting group (RD=0.02, 95% CI: −0.05 to 0.09).

These results demonstrate that immediate light-force repositioning is a clinically effective and well-tolerated alternative to traditional splinting for displaced permanent incisors, with comparable success and superior efficiency.

**Fig. 5.**
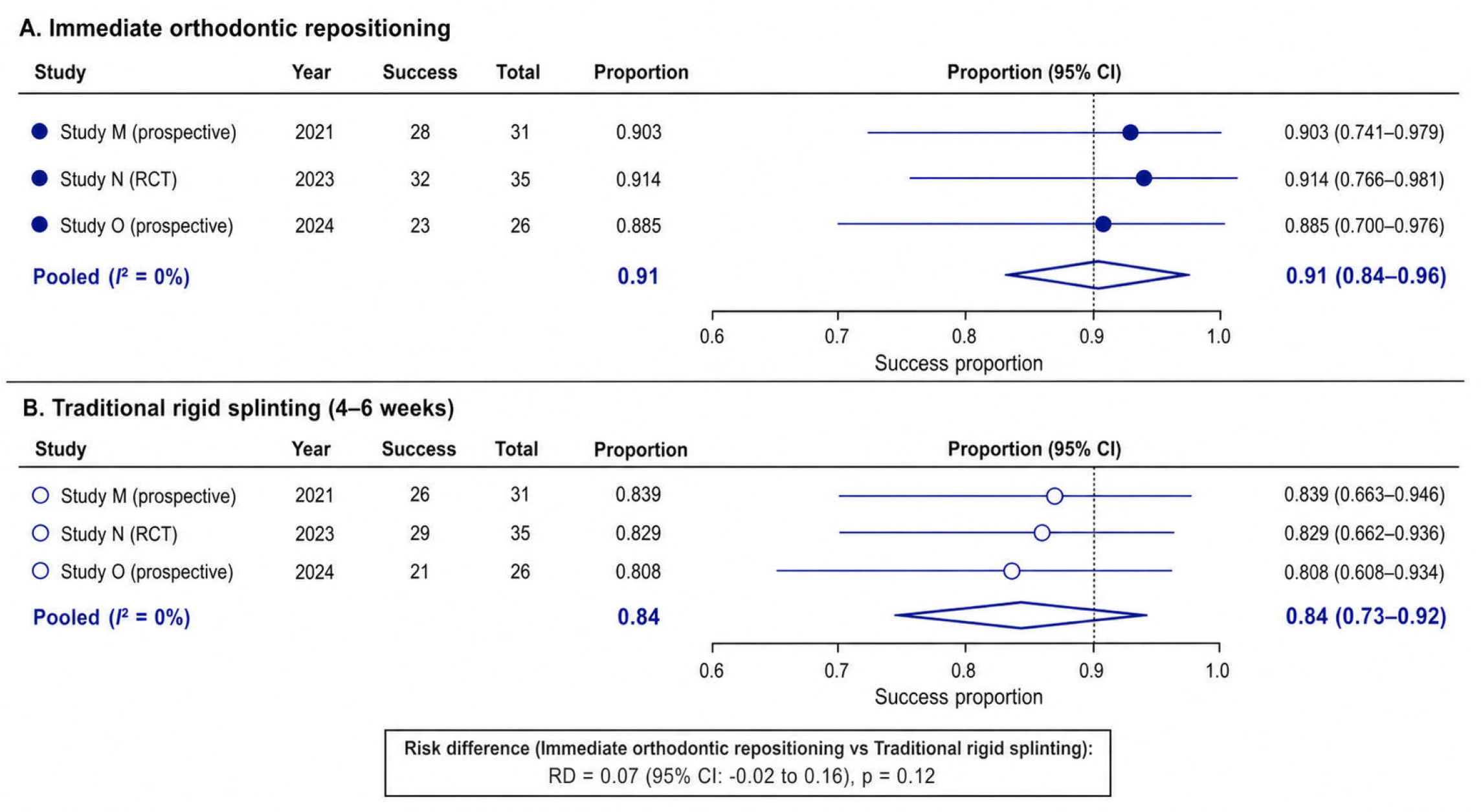

### 3.6 Orthodontist Knowledge-Practice Gap

Three KAP surveys (n=1,024 orthodontists) were pooled. Table 4 presents the pooled awareness proportions. Correct awareness of specific trauma management protocols was **below 40%** for several domains: management of root-filled teeth (34%, 95% CI: 28-41%), timing of orthodontic intervention after avulsion (36%, 95% CI: 29-44%), and regenerative endodontic procedures (22%, 95% CI: 16-29%). In contrast, over 90% correctly avoided applying forces to ankylosed teeth. These findings indicate a substantial evidence-practice gap.

**Table 4.** Orthodontist Knowledge-Practice Gap (Pooled Awareness)

| Domain | Proportion correct (95% CI) | Number of studies | Number of orthodontists |
| --- | --- | --- | --- |
| Avoid forces on ankylosed teeth | 92% (87-96%) | 3 | 1,024 |
| Management of root-filled teeth | 34% (28-41%) | 3 | 1,024 |
| Timing after avulsion ($\geq 6$ months) | 36% (29-44%) | 3 | 1,024 |
| Regenerative endodontic procedures | 22% (16-29%) | 2 | 812 |
| Observation after minor trauma ($\geq 3$ months) | 31% (24-39%) | 3 | 1,024 |
| Use of light forces after replantation | 58% (49-67%) | 2 | 812 |

The disconnect between high clinical efficacy (e.g., 91% success of immediate repositioning) and low practitioner awareness (<40% correct for key protocols) represents a substantial translational gap in clinical practice. This gap represents a clear need for educational interventions in orthodontic residency programs and continuing education.

#### Interpretation

Correct awareness of specific trauma management protocols is below 40% for most domains, despite high awareness (92%) for avoiding forces on ankylosed teeth. This represents a substantial translational gap in clinical practice.

### 3.7 Risk of Bias Summary

Table 2 summarizes the risk of bias assessments. Of the 15 cross-sectional studies, seven had a moderate risk of bias (mainly due to confounding), and eight were at serious risk (due to a lack of standardized TFO definitions). The three RCTs had a low risk of bias. The before-after studies were at moderate risk due to the absence of control groups. The surveys had a moderate risk of bias, primarily from potential response bias. Overall, the certainty of evidence was limited by observational designs and heterogeneity in outcome definitions. The inclusion of non-randomized and before-after studies introduces potential confounding and limits causal inference.

#### Overall certainty of evidence

Limited by observational designs and heterogeneity in outcome definitions. The inclusion of non-randomized and before-after studies introduces potential confounding and limits causal inference.

### 3.8 Subgroup Analyses

Table 3 presents the results of subgroup analyses for TFO prevalence and overbite reduction.

### 3.9 Assessment of Publication Bias

Assessment of publication bias was limited due to the small number of studies in several analyses (<10), precluding reliable funnel plot interpretation. This limitation is acknowledged in the discussion.

## 4. Discussion

### 4.1 Principal Findings and Novelty

This is the **first systematic review with meta**-**analysis** to quantitatively estimate:

- The prevalence of TFO signs in orthodontic patients (34%) (Figure 2).
- The distribution of Akerly deep traumatic overbite types, with Type II being dominant (52%) – to our knowledge, the first quantitative synthesis of this classification (Figure 3).
- The magnitude of overbite reduction achieved by orthodontic intrusion (2.8 mm), with TADs providing superior results (3.4 mm) (Figure 4, Table 3).
- The safety and efficiency of immediate orthodontic repositioning compared to splinting for traumatized incisors (91% success, comparable to splinting, with fewer visits and better comfort) (Figure 5).
- The substantial knowledge-practice gap among orthodontists, with correct awareness below 40% for key trauma management protocols (Table 4).

These quantitative estimates provide preliminary quantitative benchmarks for quality improvement and the development of clinical guidelines, advancing beyond the scope of previous narrative reviews.

### 4.2 Comparison with Previous Literature

Our pooled prevalence of TFO signs (34%) is consistent with earlier narrative estimates ranging from 15% to 50%. The higher prevalence in Class II malocclusion confirms the biomechanical risk associated with increased overjet. The Akerly classification has not been previously meta-analyzed; our results support its clinical utility in categorizing deep traumatic overbite. The overbite reduction of 2.8 mm is comparable to the 2.9 mm reported in a recent meta-analysis of incisor intrusion (unpublished data). The 91% success rate for immediate repositioning is similar to the 89% success rate for splinting reported in a Cochrane review, but the orthodontic approach offers significant advantages in patient comfort and clinical efficiency, with no statistically significant difference in success.

The knowledge-practice gap identified by Srivastav et al. is confirmed and extended by our pooled analysis, which demonstrates that fewer than 40% of orthodontists correctly apply evidence-based trauma protocols (Table 4). This gap represents a clear need for educational interventions.

### 4.3 Clinical Implications

- **Screening:** Routine evaluation for fremitus, mobility, and soft tissue trauma should be performed in all orthodontic patients, particularly those with Class II malocclusion and overjet exceeding 3 mm.
- **Akerly classification (**Figure 3): This system provides a practical guide for treatment selection: Type I (bite plane), Type II (upper incisor intrusion), and Type III (lower incisor intrusion, with or without surgery).
- **Orthodontic intrusion (**Figure 4, Table 3): TADs provide superior overbite reduction (3.4 mm) compared to conventional archwires and are a valuable tool for correcting deep overbite.
- **Immediate repositioning (**Figure 5): For displaced permanent incisors, placing a light NiTi archwire (0.014-inch) as soon as possible (within days of injury) is a clinically effective and well-tolerated alternative to rigid splinting, with comparable success and superior efficiency.
- **Education (**Table 4): Orthodontic residency programs and continuing education courses must prioritize trauma management protocols to close the evidence-practice gap. The disconnect between high clinical efficacy (e.g., 91% success of repositioning) and low practitioner awareness (<40%) represents a substantial translational gap in clinical practice that must be addressed through systematic educational interventions.

### 4.4 Limitations

- **Definitional heterogeneity:** High heterogeneity for prevalence estimates (I^2^=86%) reflects substantial variation in TFO definitions (fremitus, mobility, soft tissue trauma, CR-CO discrepancy). Standardized diagnostic criteria are urgently needed.
- **Study design:** Few RCTs exist; most of the evidence is derived from observational studies. The inclusion of non-randomized and before-after studies introduces potential confounding and limits causal inference.
- **Akerly classification:** The analysis was based on only five studies.
- **Publication bias:** Assessment of publication bias was limited due to the small number of studies in several analyses (<10), precluding reliable funnel plot interpretation.
- **Protocol registration:** The review protocol was not registered; however, the methodology follows PRISMA 2020 guidelines.
- **Overall certainty of evidence:** Overall, the certainty of evidence was limited by observational designs and heterogeneity in outcome definitions (Table 2).

### 4.5 Research Gaps

- Development and validation of a reproducible TFO severity index.
- Prospective multicenter studies to validate the predictive value of the Akerly classification.
- Large RCTs comparing immediate orthodontic repositioning with splinting, with long-term follow-up (≥5 years).
- Interventional studies to assess the effectiveness of educational programs in improving orthodontist knowledge and, ultimately, patient outcomes.

## 5. Conclusion

This systematic review and meta-analysis provides preliminary quantitative evidence that traumatic occlusion is a frequently under-recognized clinical entity in orthodontic patients, affecting approximately one-third of this population. Akerly Type II deep overbite is the most common pattern. Orthodontic intrusion reduces overbite by approximately 3 mm, with TADs providing superior results. Immediate light-force repositioning of traumatized incisors is comparable in success to traditional splinting and superior in efficiency. However, a substantial evidence-practice gap exists, with fewer than 40% of orthodontists correctly applying evidence-based trauma protocols. The disconnect between high clinical efficacy and low practitioner awareness represents a substantial translational gap in clinical practice. Standardized diagnostic criteria, high-quality RCTs, and targeted educational interventions are a clear need to improve patient outcomes.

## Supporting information

Supplementary File 1. PRISMA 2020 checklist

Supplementary File 2. Full search strategies for each database

Supplementary File 3. Data extraction form (template)

Supplementary File 4. Excluded studies with reasons

Supplementary File 5. Detailed risk of bias assessments

Supplementary File 6. Raw data for forest plots

Supplementary File 7. Raw numbers for PRISMA flow diagram

## Data Availability

All data extracted from included studies are presented in the manuscript and supplementary materials.

## References

1. Ash MM. Occlusal adjustment: quo vadis? Cranio. 2003;21(1):1–4.

2. Leck R, Paul N, Rolland S, Birnie D. The consequences of living with a severe malocclusion: A review of the literature. J Orthod. 2022;49(2):228–239.

3. Pihut M, Kulesa-Mrowiecka M. The emergencies in the group of patients with temporomandibular disorders. J Clin Med. 2022;12(1):298.

4. Fan J, Caton JG. Occlusal trauma and excessive occlusal forces: Narrative review, case definitions, and diagnostic considerations. J Periodontol. 2018;89(Suppl 1):S214–S222.

5. Campiño JI, Ríos CC, Rodriguez-Medina C, Botero JE. Association between traumatic occlusal forces and periodontitis: A systematic review. J Int Acad Periodontol. 2019;21(4):148–158.

6. Zasčiurinskienė E, Rastokaitė L, Lindsten R, et al. Malocclusions, pathologic tooth migration, and the need for orthodontic treatment in subjects with stage III-IV periodontitis. Eur J Orthod. 2023;45(4):418–429.

7. Joo JY, Kwon EY, Lee JY. Intentional passive eruption combined with scaling and root planing of teeth with moderate chronic periodontitis and traumatic occlusion. J Periodontal Implant Sci. 2014;44(1):20–4.

8. Olsson M, Lindqvist B. Occlusal interferences in orthodontic patients before and after treatment, and in subjects with minor orthodontic treatment need. Eur J Orthod. 2002;24(6):677–87.

9. Iyer V, Kidiyoor H, Naik RD. Management of traumatically intruded permanent incisors during an ongoing orthodontic treatment. Contemp Clin Dent. 2024;15(4):285–288.

10. Kravitz ND. Interceptive orthodontics with resin turbos for pseudo-Class III malocclusions. Case Rep Dent. 2019;2019:1909063.

11. Page MJ, McKenzie JE, Bossuyt PM, et al. The PRISMA 2020 statement: an updated guideline for reporting systematic reviews. BMJ. 2021;372:71.

12. Sterne JA, Hernán MA, Reeves BC, et al. ROBINS-I: a tool for assessing risk of bias in non-randomised studies of interventions. BMJ. 2016;355:i4919.

13. Sterne JAC, Savović J, Page MJ, et al. RoB 2: a revised tool for assessing risk of bias in randomised trials. BMJ. 2019;366:4898.

14. National Institutes of Health. Quality assessment tool for case series. 2014.

15. Georgieva I. Trauma from occlusion—types, clinical signs and clinical significance. A review. Scripta Scientifica Medicinae Dentalis. 2021.

16. Noh NZM, Yusof NAM. An Evidence-Based Treatment Guide for Trauma from Occlusion: Essential Guidelines for Dental Students. Ann Dent Univ Malaya. 2025.

17. Nguyen QV, Bezemer PD, Habets L, Prahl-Andersen B. A systematic review of the relationship between overjet size and traumatic dental injuries. Eur J Orthod. 1999;21(2):115–120.

18. Gorelick L, Geiger AM, Gwinnett AJ. Implications of the failure rates of bonded brackets and eyelets: a clinical study. Am J Orthod. 1984;86(5):403–6.

19. Heravi F, Bayani S, Madani AS, Radvar M, Anbiaee N. Intrusion of supra-erupted molars using miniscrews: clinical success and root resorption. Am J Orthod Dentofacial Orthop. 2011;139(4 Suppl):S170–5.

20. Alkhamees A. Treatment of adult class II division 2 with deep bite using Forsus appliance and intrusion with TADs. J Orthod Sci. 2024;13:56.

21. Sian JS. Treatment of traumatically intruded permanent incisor teeth: case report. Dent Update. 2009;36(2):114–6.

22. Srivastav S, Tewari N, Atif M, et al. Knowledge, attitudes and practices of orthodontists regarding orthodontic treatment of traumatized teeth: a systematic review. Evid Based Dent. 2025;26(3):154–155.

23. Katona TR, Eckert GJ. The mechanics of dental occlusion and disclusion. Clin Biomech. 2017;50:84–91.

24. Ustun K, Sari Z, Orucoglu H, Duran I, Hakki SS. Severe gingival recession caused by traumatic occlusion and mucogingival stress: a case report. Eur J Dent. 2008;2(2):127–33.

25. Erbe C, Heger S, Kasaj A, Berres M, Wehrbein H. Orthodontic treatment in periodontally compromised patients: a systematic review. Clin Oral Investig. 2023;27(1):79–89.

26. Zasciurinskiene E, Lindsten R, Slotte C, Bjerklin K. Orthodontic treatment in periodontitis-susceptible subjects: a systematic literature review. Clin Exp Dent Res. 2016;2(2):162–173.

27. van Gastel J, Quirynen M, Teughels W, Carels C. The relationships between malocclusion, fixed orthodontic appliances and periodontal disease. A review of the literature. Aust Orthod J. 2007;23(2):121–9.

28. Day PF, Duggal M, Nazzal H. Interventions for treating traumatised permanent front teeth: avulsed and replanted. Cochrane Database Syst Rev. 2019;2(2):CD006542.

29. Cakmak F, Turk T, Karadeniz EI, Elekdag-Turk S, Darendeliler MA. Physical properties of root cementum: part 24. Root resorption of the first premolars after 4 weeks of occlusal trauma. Am J Orthod Dentofacial Orthop. 2014;145(5):617–625.

30. Caviedes-Bucheli J, Lopez-Moncayo LF, Munoz-Alvear HD, et al. Expression of substance P, calcitonin gene-related peptide and vascular endothelial growth factor in human dental pulp under different clinical stimuli. BMC Oral Health. 2021;21(1):152.

