## Supplementary File 1. PRISMA 2020 checklist for "Traumatic Occlusion in Orthodontics: A Systematic Review and Meta-Analysis of Prevalence, Classification, Treatment Outcomes, and the Evidence–Practice Gap"

| Section and Topic | Item # | Checklist item | Location where item is reported |
| --- | --- | --- | --- |
| **TITLE** |  |  |  |
| Title | 1 | Identify the report as a systematic review. | Title page |
| **ABSTRACT** |  |  |  |
| Abstract | 2 | See PRISMA 2020 for Abstracts checklist. | Abstract |
| **INTRODUCTION** |  |  |  |
| Rationale | 3 | Describe the rationale for the review in the context of existing knowledge. | Introduction |
| Objectives | 4 | Provide an explicit statement of the objective(s) or question(s) the review addresses. | Introduction (end) |
| **METHODS** |  |  |  |
| Eligibility criteria | 5 | Specify the inclusion and exclusion criteria for the review. | Section 2.2 |
| Information sources | 6 | Specify all databases, registers, websites, organisations, and other sources searched. | Section 2.1 |
| Search strategy | 7 | Present the full search strategies for all databases and registers. | Section 2.1 (full string provided) |
| Selection process | 8 | Specify the methods used to decide whether a study met the inclusion criteria. | Section 2.3 |
| Data collection process | 9 | Describe the methods used to collect data from reports. | Section 2.3 |
| Data items | 10 | List and define all outcomes for which data were sought. | Section 2.3 |
| Study risk of bias assessment | 11 | Specify the methods used to assess risk of bias. | Section 2.4 |
| Effect measures | 12 | Specify for each outcome the effect measure(s) (e.g., risk ratio, mean difference). | Section 2.5 |
| Synthesis methods | 13 | Describe the methods used to synthesise results. | Section 2.5 |
| Reporting bias assessment | 14 | Describe any methods used to assess risk of bias due to missing results. | Section 2.5 |
| Certainty assessment | 15 | Describe any methods used to assess certainty (or confidence) in the body of evidence. | Section 2.5 |
| **RESULTS** |  |  |  |
| Study selection | 16 | Describe the results of the search and selection process. | Section 3.1, Figure 1 |
| Study characteristics | 17 | Cite each included study and present its characteristics. | Table 1 |
| Risk of bias in studies | 18 | Present assessments of risk of bias for each included study. | Table 2 |
| Results of individual studies | 19 | For all outcomes, present for each study: summary statistic and risk of bias. | Figures 2‑5 |
| Results of syntheses | 20 | For each meta‑analysis, present summary estimate and measures of heterogeneity. | Figures 2‑5 |
| Reporting biases | 21 | Present assessments of risk of bias due to missing results. | Section 3.9 |
| Certainty of evidence | 22 | Present assessments of certainty (or confidence) in the body of evidence. | Table 2 |
| **DISCUSSION** |  |  |  |
| Discussion | 23 | Provide a general interpretation of the results. | Section 4 |
| **OTHER** |  |  |  |
| Registration | 24 | Provide registration information for the review. | Not registered (justification provided in Methods) |
| Conflict of interest | 25 | Declare any competing interests. | Title page |
| Funding | 26 | Report sources of funding. | Title page |
| Data availability | 27 | Report availability of data. | Title page |
