## Supplementary File 2. Full search strategies for each database for "Traumatic Occlusion in Orthodontics: A Systematic Review and Meta-Analysis of Prevalence, Classification, Treatment Outcomes, and the Evidence–Practice Gap"

### **PubMed (MEDLINE)**

**Search date:** December 2025
**Search string:**

("traumatic occlusion" OR "occlusal trauma" OR "traumatic overbite" OR "Akerly classification") AND ("orthodontics" OR "intrusion" OR "repositioning" OR "pathologic migration" OR "overjet")

**Filters applied:** Publication date from 2000 to 2025
**Results:** 57

### **PubMed Central (PMC)**

**Search date:** December 2025
**Search string:**

("traumatic occlusion" OR "occlusal trauma" OR "traumatic overbite" OR "traumatic malocclusion")

**Filters applied:** Publication date from 2000 to 2025; Full‑text available
**Results:** 538

### **Cochrane Library**

**Search date:** December 2025
**Search string:**

("traumatic occlusion" OR "occlusal trauma" OR "traumatic overbite" OR "Akerly classification") AND ("orthodontics" OR "intrusion" OR "repositioning" OR "pathologic migration" OR "overjet")

**Components searched:** Cochrane Reviews, CENTRAL trials
**Results:** 11 total (2 reviews, 9 trials)

**Cochrane Reviews retrieved:**

1. Millett DT, et al. Orthodontic treatment for deep bite and retroclined upper front teeth in children. Cochrane Database Syst Rev. 2018.
2. Ye Z, et al. Periodontal therapy for primary or secondary prevention of cardiovascular disease in people with periodontitis. Cochrane Database Syst Rev. 2022.

**CENTRAL trials retrieved:**

1. Gorelick L, Geiger AM, Gwinnett AJ. Implications of the failure rates of bonded brackets and eyelets: a clinical study. Am J Orthod. 1984;86(5):403-406.
2. Cakmak F, et al. Physical properties of root cementum: part 24. Root resorption of the first premolars after 4 weeks of occlusal trauma. Am J Orthod Dentofacial Orthop. 2014;145(5):617-625.
3. Caviedes-Bucheli J, et al. Expression of substance P, calcitonin gene-related peptide and vascular endothelial growth factor in human dental pulp under different clinical stimuli. BMC Oral Health. 2021;21(1):152.
4. IRCT2017092036203N2. The effect of laser therapy on the healing of human palatal wound after free gingival graft surgery. 2017.
5. (plus 5 additional trial registry records)

### **Google Scholar**

**Search date:** December 2025
**Search string:**

("traumatic occlusion" OR "occlusal trauma" OR "traumatic overbite")

**Filters applied:** Publication date from 2000 to 2025; sorted by relevance
**Limitation:** First 200 results screened
**Results:** Approximately 3,930 (only first 200 reviewed)
