## Supplementary File 3. Data extraction form (template) for "Traumatic Occlusion in Orthodontics: A Systematic Review and Meta-Analysis of Prevalence, Classification, Treatment Outcomes, and the Evidence–Practice Gap"

| Field | Data extracted |
| --- | --- |
| **Study identification** |  |
| Author(s) |  |
| Year of publication |  |
| Country |  |
| **Study characteristics** |  |
| Study design |  |
| Sample size (n) |  |
| Age range (years) |  |
| Male/Female ratio |  |
| Malocclusion type |  |
| **TFO definition** |  |
| Fremitus (yes/no) |  |
| Mobility (yes/no) |  |
| Soft tissue trauma (yes/no) |  |
| CR‑CO discrepancy (yes/no) |  |
| Occlusal interference (yes/no) |  |
| **Akerly classification (if applicable)** |  |
| Type I |  |
| Type II |  |
| Type III |  |
| **Intervention (if applicable)** |  |
| Type |  |
| Force (grams) |  |
| Duration (weeks/months) |  |
| **Outcomes** |  |
| Prevalence proportion |  |
| Overbite reduction (mm) |  |
| Success rate (%) |  |
| Root resorption (any) |  |
| Follow‑up duration |  |
| **Risk of bias** |  |
| ROBINS‑I / RoB 2 / NIH score |  |
