## Supplementary File 4. Excluded studies with reasons for "Traumatic Occlusion in Orthodontics: A Systematic Review and Meta-Analysis of Prevalence, Classification, Treatment Outcomes, and the Evidence–Practice Gap"

| Study | Reason for exclusion |
| --- | --- |
| Amaral MF, et al. (2017) | Animal study |
| Devi S, Sundari S (2023) | No original data (abstract only) |
| Georgieva I (2021) | Narrative review, no original data |
| Fan J, Caton JG (2018) | Narrative review, no original data |
| Noh NZM, Yusof NAM (2025) | Educational guide, no original data |
| Wang Y, et al. (2025) | Animal study (rat model) |
| Katona TR, Eckert GJ (2017) | Experimental biomechanics, no clinical outcomes |
| (plus 53 additional records) | Not relevant to TFO/orthodontics; animal studies; case reports n<5; reviews; implant/prosthesis only |

**Total excluded:** 60
