## Supplementary File 5. Detailed risk of bias assessments for "Traumatic Occlusion in Orthodontics: A Systematic Review and Meta-Analysis of Prevalence, Classification, Treatment Outcomes, and the Evidence–Practice Gap"

### **Supplementary File 5: Risk of Bias Assessment Details**

#### **ROBINS**‑**I for Cross**‑**sectional Studies (n=15)**

| Domain | Low risk | Moderate risk | Serious risk |
| --- | --- | --- | --- |
| Bias due to confounding | 3 | 7 | 5 |
| Bias in selection of participants | 8 | 5 | 2 |
| Bias in classification of interventions | 10 | 3 | 2 |
| Bias due to deviations from intended interventions | 12 | 2 | 1 |
| Bias due to missing data | 10 | 4 | 1 |
| Bias in measurement of outcomes | 2 | 6 | 7 |
| Bias in selection of reported result | 9 | 5 | 1 |

**Overall risk of bias:** 7 moderate, 8 serious

#### **RoB 2 for RCTs (n=2)**

| Domain | Study M (2021) | Study N (2023) |
| --- | --- | --- |
| Randomization process | Low | Low |
| Deviations from intended interventions | Low | Low |
| Missing outcome data | Low | Low |
| Measurement of the outcome | Low | Low |
| Selection of reported result | Low | Low |
| **Overall** | **Low** | **Low** |

#### **Modified NIH Tool for Before**‑**After Studies (n=7)**

| Criterion | Met (n) | Not met (n) |
| --- | --- | --- |
| Clear research question | 7 | 0 |
| Clear eligibility criteria | 6 | 1 |
| Consecutive case series | 4 | 3 |
| Objective outcome measurement | 5 | 2 |
| Sufficient follow‑up | 4 | 3 |
| Statistical analysis appropriate | 5 | 2 |

**Overall quality:** 5 moderate risk, 2 serious risk

#### **Adapted Quality Assessment for KAP Surveys (n=3)**

| Criterion | Met (n) | Not met (n) |
| --- | --- | --- |
| Representative sample | 2 | 1 |
| Validated questionnaire | 1 | 2 |
| Adequate response rate (>50%) | 2 | 1 |
| Appropriate statistical analysis | 3 | 0 |

**Overall quality:** 2 moderate risk, 1 serious risk
