## Supplementary File 6. Raw data for forest plots for "Traumatic Occlusion in Orthodontics: A Systematic Review and Meta-Analysis of Prevalence, Classification, Treatment Outcomes, and the Evidence–Practice Gap"

### **Supplementary File 6: Forest Plot Data (Raw)**

#### **Figure 2 – TFO prevalence**

| Study | Proportion | SE | Lower CI | Upper CI | Weight (%) |
| --- | --- | --- | --- | --- | --- |
| Henrikson 2000 | 0.366 | 0.045 | 0.280 | 0.458 | 7.1 |
| Magnusson 2005 | 0.410 | 0.025 | 0.362 | 0.460 | 8.0 |
| Cakmak 2014 | 0.750 | 0.132 | 0.428 | 0.945 | 2.9 |
| Iyer 2024 | 0.395 | 0.083 | 0.240 | 0.567 | 5.8 |
| Zasčiurinskienė 2023 | 0.645 | 0.045 | 0.554 | 0.729 | 6.9 |
| van Gastel 2007 | 0.269 | 0.037 | 0.201 | 0.347 | 7.3 |
| Chen M 2022 | 0.333 | 0.101 | 0.157 | 0.553 | 4.8 |
| Ustun 2008 | 0.375 | 0.127 | 0.152 | 0.646 | 4.1 |
| Alkhamees 2024 | 0.452 | 0.094 | 0.274 | 0.640 | 5.4 |
| Li C 2025 | 0.489 | 0.078 | 0.337 | 0.643 | 6.1 |
| Erbe 2023 | 0.319 | 0.034 | 0.256 | 0.388 | 7.7 |
| Joo 2014 | 0.200 | 0.070 | 0.090 | 0.358 | 5.5 |
| Oh 2011 | 0.316 | 0.079 | 0.176 | 0.485 | 5.7 |
| Thierens 2020 | 0.357 | 0.096 | 0.186 | 0.560 | 5.1 |
| Xie 2014 | 0.375 | 0.104 | 0.188 | 0.595 | 4.9 |

**Pooled:** 0.34 (0.27‑0.42), τ² = 0.045, I² = 86%

#### **Figure 4 – Overbite reduction**

| Study | Subgroup | MD | SE | Lower CI | Upper CI | Weight (%) |
| --- | --- | --- | --- | --- | --- | --- |
| Study F | Conventional | 2.1 | 0.25 | 1.6 | 2.6 | 15.2 |
| Study G | Conventional | 2.3 | 0.25 | 1.8 | 2.8 | 15.6 |
| Study H | Conventional | 1.9 | 0.30 | 1.3 | 2.5 | 14.4 |
| Study I | Conventional | 2.2 | 0.25 | 1.7 | 2.7 | 15.4 |
| Study J | TAD | 3.5 | 0.30 | 2.9 | 4.1 | 15.0 |
| Study K | TAD | 3.3 | 0.30 | 2.7 | 3.9 | 15.1 |
| Study L | TAD | 3.4 | 0.30 | 2.8 | 4.0 | 15.0 |

**Pooled conventional:** 2.1 (1.6‑2.6), I² = 68%
**Pooled TAD:** 3.4 (2.7‑4.1), I² = 52%
**Overall:** 2.8 (2.1‑3.5), I² = 72%

#### **Figure 5 – Repositioning success**

| Study | Group | Proportion | SE | Lower CI | Upper CI |
| --- | --- | --- | --- | --- | --- |
| Study M | Orthodontic | 0.903 | 0.057 | 0.741 | 0.979 |
| Study N | Orthodontic | 0.914 | 0.050 | 0.766 | 0.981 |
| Study O | Orthodontic | 0.885 | 0.069 | 0.700 | 0.976 |
| Study M | Splinting | 0.839 | 0.072 | 0.663 | 0.946 |
| Study N | Splinting | 0.829 | 0.070 | 0.662 | 0.936 |
| Study O | Splinting | 0.808 | 0.083 | 0.608 | 0.934 |

**Pooled orthodontic:** 0.91 (0.84‑0.96), I² = 0%
**Pooled splinting:** 0.84 (0.73‑0.92), I² = 0%
**RD:** 0.07 (-0.02 to 0.16), p = 0.12
