## Supplementary File 7. Raw numbers for PRISMA flow diagram for "Traumatic Occlusion in Orthodontics: A Systematic Review and Meta-Analysis of Prevalence, Classification, Treatment Outcomes, and the Evidence–Practice Gap"

### **Supplementary File 7: PRISMA Flow Diagram Data (Raw)**

| Stage | Number |
| --- | --- |
| PubMed | 57 |
| PubMed Central | 538 |
| Cochrane Library | 11 |
| Google Scholar | 3,930 |
| **Total identified** | **4,536** |
| After duplicates removed | 3,214 |
| Records screened | 3,214 |
| Records excluded | 3,127 |
| Full‑text assessed | 87 |
| Full‑text excluded | 60 |
| **Studies included** | **27** |
